# Genetic risk estimates for offspring of patients with Stargardt disease

**DOI:** 10.1101/2021.08.11.21261888

**Authors:** Stéphanie S. Cornelis, Esmee H. Runhart, Miriam Bauwens, Zelia Corradi, Elfride de Baere, Susanne Roosing, Lonneke Haer-Wigman, Claire-Marie Dhaenens, Anneke T. Vulto-van Silfhout, Frans P.M. Cremers

## Abstract

**BACKGROUND:** Genetic counseling in autosomal recessive Stargardt disease (STGD1) is complicated because of unknown frequencies of pathogenic *ABCA4* alleles across populations, variable and unknown severity of *ABCA4* alleles, and incomplete penetrance.

**METHODS:** In this cross-sectional study, published *ABCA4* variants were categorized by severity based on previous functional and clinical studies and current statistical comparisons of their frequencies in patients versus the general population, their observed versus expected homozygous occurrence in patients, and their occurrence in combination with established mild alleles in patients. The sum allele frequencies of these severity categories were used to estimate inheritance risks for offspring of STGD1 patients and carriers of pathogenic *ABCA4* variants.

**RESULTS:** The risk for offspring of a STGD1 patient with the ‘severe|severe’ genotype or a ‘severe|mild with complete penetrance’ genotype to develop STGD1 at some moment in life was estimated at 2.8-3.1% (1 in 35-32 individuals) and 1.6-1.8% (1 in 62-57 individuals), respectively. The risk to develop STGD1 in childhood was estimated to be 2 to 4-fold lower: 0.7-0.8% (1 in 148-124) and 0.3-0.4% (1 in 295-248), respectively. For offspring of an unaffected *ABCA4* variant carrier from a STGD1 family who carries one severe or one mild *ABCA4* variant with complete penetrance, the risk to develop STGD1 throughout life is 1.4-1.6% (1 in 71-64) and 0.19-0.21% (1 in 516-487), respectively.

**CONCLUSION:** We propose a genotype-based personalized counseling approach to appreciate the large differences in inheritance risk between individuals. We advocate considering the lower risk of early-onset STGD1 compared with the total STGD1 risk.

## Introduction

Autosomal recessive Stargardt disease due to bi-allelic variants in *ABCA4* (STGD1; MIM 248200) represents the most prevalent inherited maculopathy, estimated to occur in 1 in 10,000 individuals.^1^ Although originally considered a juvenile macular degeneration, patients may experience initial visual complaints between the first and the eighth decade of life.^2-7^ STGD1 results in visual impairment due to central or pericentral vision loss, impaired color vision, distorted vision and/or visual field defects, with legal blindness after a median of 12 disease years.^3,8,9^ Currently, in the absence of a treatment for STGD1, a major part of its management involves counseling about prognosis e.g. to help make career choices, and inheritance risk e.g. to aid in family planning.

The risk of passing the disease to (future) children is a common concern of STGD1 patients and their relatives. This concern needs careful consideration because of the high frequency of pathogenic *ABCA4* variants in the general population. High *ABCA4* carrier frequencies are illustrated by common observations of pseudodominant inheritance,^10-14^ and different combinations of disease-causing *ABCA4* variants among siblings.^9,15^

The large difference in the age of onset of STGD1 between patients is hypothesized to be mainly caused by the variable amount of residual ABCA4 activity. The combination of *ABCA4* variants of different severity, ranging from null (severe) alleles, moderately severe alleles, to mild alleles, influences the clinical expression.^13,16,17^ Especially relevant to inheritance risk is that combinations of pathogenic *ABCA4* variants may or may not cause STGD1 depending on the variant severity. Two mild alleles, in principle, do not cause STGD1. Another layer of complexity was added to the existing *ABCA4* genotype-phenotype correlation model, and thus genetic counseling, by evidence of incomplete penetrance.^18,19^ We calculated that the very frequent hypomorphic variant c.5603A>T (p.(Asn1868Ile)) (allele frequency (AF) of 4.2% in the Genome Aggregation Database (gnomAD)), when in *trans* with a null allele, has a very low penetrance (<5%).^9,20^ Moreover, a female predilection among patients was observed for this variant and for another frequent variant, c.5882G>A (p.Gly1961Glu).^9,21^ These findings led to the hypothesis that ∼25% of STGD1 patients display multifactorial or polygenic inheritance in which two *ABCA4* alleles are a prerequisite to develop STGD1 but additional genetic or non-genetic modifiers play a significant role.

The *ABCA4* mutational landscape has further been expanded by the discovery of causative deep-intronic variants and structural variants. Targeted whole gene sequencing has revealed 42 causal deep-intronic variants with variable effects on RNA splicing in ∼5% of *ABCA4* alleles and ∼10% of STGD1 cases.^22-27^ Moreover, 46 different structural variants have been identified in ∼2% of STGD1 cases.^24-26^ Finally, at least 53 complex *ABCA4* alleles have been identified, consisting of multiple disease-causing variants in *cis*,^25,28^ illustrating the importance of establishing the phase of the variants found in diagnostic studies.

Despite the advances in genetic testing and increased knowledge of pathogenic variants, most variants are of unknown severity.^4^ As shown by the proposed *ABCA4* genotype-phenotype correlation model, knowledge of not only the pathogenicity but also the severity of *ABCA4* variants is crucial in estimating inheritance risk. Existing guidelines for the interpretation of pathogenicity of variants are not tailored to assess variant severity. The commonly used 5-tier American College of Medical Genetics and Genomics and the Association for Molecular Pathology (ACMG-AMP) system addresses the likelihood of a variant’s pathogenicity on a scale from benign (1) to causal (5) which in no way relates to its severity.^29^ For instance, the frequent mild *ABCA4* variant c.5882A>G (p.(Gly1961Glu)) is considered ‘likely pathogenic’ according to the ACMG-AMP guidelines, whereas variants classified as variants of unknown significance by ACMG-AMP may well be severe. Therefore, genetic counseling of STGD1 families demands a genotype-based approach that considers variant severity.

In this study, we aimed to calculate the risk for offspring of STGD1 patients and carriers to develop STGD1. We used published *ABCA4* genotype data from patients and *ABCA4* AF data from the general population to assess the severity of 1,619 *ABCA4* variants. The sum frequencies of *ABCA4* alleles of different severities were calculated in the general population ethnically matched to the patient population. Using Hardy-Weinberg’s principle, inheritance risk was assessed for offspring of STGD1 patients and *ABCA4* variant carriers.

## Methods

Currently non-existing terminology limits the use of inclusive language. By ‘offspring’ and ‘child’ we refer to the genetic offspring and child, and by ‘partner’ we refer to the other genetic parent of the offspring. Similarly, we use the term ‘ethnicity’ to refer to groups of people who share a certain degree of genetic similarity; not to refer to cultural elements. We stress that we distance ourselves from problematic use of the term ethnicity as it is sometimes used to describe ‘normal’ and ‘other’ groups or people.

### 1. Dataset compositions: the bi-allelic patient (BAP) dataset and the ethnically-matched gnomAD (EM-gnomAD) dataset

Inheritance risk calculations require knowledge of all *ABCA4* variants that contribute to the disease in the given population, i.e. which variants are disease-causing and how severe are those variants. Furthermore, the inheritance risk calculations require knowledge of the sum AF of each severity category in the general population ethnically matched to the patient population under study. In order to obtain this information, a patient and a control dataset must be established.

#### Selection of the BAP dataset

We collected all publications until January 1^st^, 2020 that contain *ABCA4* variants in patients with an autosomal recessive retinal dystrophy (Supplemental Materials and Methods). Reported variants in bi-allelic patients were collected per patient. Only bi-allelic patients were selected because mono-allelic patients are less likely to have *ABCA4* associated retinopathy. Patient records with the same variants that had been published by overlapping authors were considered duplicates and were removed from the dataset when there was no conflicting data on gender, ethnicity or age at onset (n=751). Furthermore, affected family members were excluded from the dataset (n=58). The resulting dataset (n=5,579) will be referred to as the bi-allelic patient dataset (BAP dataset).

#### Composition of the EM-gnomAD dataset

*ABCA4* AF data were derived from the Genome Aggregation Database (gnomAD), downloaded on 13 April 2021 and were used to establish a control dataset that ethnically matches the BAP dataset: the ethnically matched-gnomAD (EM-gnomAD) dataset (Supplemental Materials and Methods).

The sum AFs of *ABCA4* alleles categorized by severity: severe, moderately severe, mild with complete penetrance (mild^CP^), mild with incomplete penetrance (mild^IP^)

### 2. The combination of test results from the following three tests were used for the severity category assignment: AF test, Homozygosity test, Severity odds ratio test

#### AF test: Benign test

To identify benign variants, we compared the frequency of each variant in the BAP dataset to the frequency of that variant in the EM-gnomAD dataset using Fisher Exact and creating an odds ratio (OR) for each variant. An OR <1 points toward a benign nature of the variant.

#### Homozygosity test: Mild variant test

Mild variants are not expected to cause STGD1 in a homozygous configuration. Therefore, a lower homozygous frequency in the BAP dataset than expected based on the AF in the general population indicates that a variant is mild. We compared the observed homozygous frequency of each variant in the BAP dataset to the expected homozygous frequency based on the AF in the EM-gnomAD dataset assuming they would cause disease in a homozygous state using Fisher Exact: Assuming a disease prevalence of 1:10,000, every person in the BAP dataset represents a population of 10,000 people. To calculate the expected homozygous occurrence of each variant in the BAP dataset, we therefore multiplied the EM-gnomAD AF squared with the inverse of the assumed STGD1 prevalence (1:10,000) and with the BAP dataset size

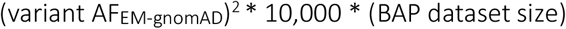

An OR was calculated for each variant. A low OR equals a low frequency of homozygous configuration and therefore points toward a mild nature of the variant.

#### Severity odds ratio: Severe variants test

Mild and moderately severe variants are not expected to cause STGD1 disease when in *trans* with a mild variant. Therefore, to distinguish severe variants from mild and moderately severe variants the ratio of each variant occurring in *trans* with previously established mild variants and previously established severe variants (Supplemental Materials and Methods) was compared to the same ratio in a reference group of previously established severe variants in the BAP dataset, using Fisher Exact. An OR was calculated for each variant. A high severity OR (>0.8) means that a variant has a relatively high frequency with mild variants and therefore points toward a severe nature of the variant, whereas a low severity OR (<0.8) points toward a mild or moderately severe nature of the variant.

#### Severity category assignment

In order to get a best estimate of sum AFs per severity category, we assigned variants to the categories ‘benign’, ‘mild^IP^’, ‘mild^CP^’, ‘moderately severe’ and ‘severe’ or multiple on the basis of the outcomes of the three aforementioned tests; the steps are described in detail in the Supplemental Materials and Methods and Supplemental Figure 1.

### 3. Sum AFs per severity category

Initial sum AFs were created per severity category. Because of the subset of variants (n=192) that were inconclusively allocated to two severity categories, we made an underestimate and an overestimate of severity AFs. In the underestimate AF, all ambiguous variants were allocated to the least severe category, while those same variants were allocated to the most severe category in the overestimate AF. The sum AFs of variants that were categorized as one of three or more categories were proportionally divided over the three pathogenic severity categories, for ‘causative of unknown severity’ and over benign, mild^CP^, moderately severe and severe for ‘not categorized’. For this, the sum AF of benign variants only included those variants with an AF <0.01 across all gnomAD populations, as we considered the more frequent benign variants as erroneously included in studies despite a high AF. The sum AF of 66 null alleles from gnomAD that were absent from the BAP dataset were added to the sum AF of the severe category. The sum AFs were further adjusted according to the factors described below.

#### Consideration of frequent complex alleles

Several frequent mild alleles occur in *cis* with other pathogenic *ABCA4* variants. Therefore, the AF was corrected for c.[769-784C>T;5603A>T], c.[1622T>C;3113C>T], c.[2588G>C;5603A>T], c.[5461-10T>C;5603A>T]. In the calculation of the sum AF of mild^IP^, variants c.769-784C>T and c.2588G>C were only partially considered (25% (Genome of The Netherlands)^30^ and 10%,^17^ respectively), because these are likely benign if c.5603A>T is not located on the same allele. Also, the AF of the severe c.5461-10T>C variant was subtracted from the AF of c.5603A>T, since they are in linkage disequilibrium.^25^

Likewise, c.3113C>T was only partially considered in the sum AF of mild^CP^ alleles, because it has a confirmed severe effect when in *cis* with c.1622C>T. Eighty-five percent of the AF of c.1622C>T was therefore subtracted from the sum AF of mild^CP^ alleles with complete penetrance, because 85% of all c.1622C>T alleles in our patient data also contains c.3113C>T and this is expected to be a similar proportion in the general population.

### 4. Inheritance risk calculations

#### *ABCA4* genotype frequencies of unaffected individuals in the EM-gnomAD dataset

We assessed the *ABCA4* genotype frequencies in the EM-gnomAD dataset, based on the Hardy-Weinberg principle (p^2^+2pq+q^2^=1). Consequently, genotypes with two alleles from the same severity category, have an occurrence of that category’s sum AF squared, while genotypes of two variants of two different categories, have an occurrence of two times the multiplication of the categories’ sum AFs. We assume that the frequencies of severe, moderately severe, mild^CP^, mild^IP^ (p.(Asn1868Ile)), and wildtype alleles add up to a total of 100%, that allele and genotype frequencies remain rather stable throughout time, that the STGD1 prevalence is equal in all analyzed populations and that genetic variation was spread evenly through the population, even though this is not the reality. Inheritance risks are calculated for situations where one of the biological parents has an unknown *ABCA4* genotype, a situation that applies to most unaffected individuals in the general population. However, some individuals with a pathogenic *ABCA4* genotype may be clinically diagnosed after their child is born and are therefore likely considered an unaffected individual at the time of risk assessment. The mean age of biological parents of children born in 2018 in the European Union was 32 years (http://appsso.eurostat.ec.europa.eu/nui/submitViewTableAction.do; https://www.cbs.nl/en-gb/news/2011/39/one-in-six-first-time-fathers-over-40). Patients with the genotypes severe|mild^CP^ and moderate|moderate are generally diagnosed with STGD1 around this age. Therefore, only the proportion of individuals with these genotypes and a diagnosis >32 age in our patient cohort (E.H.R., personal communication) was taken into consideration in the calculations of inheritance risk involving an unaffected parent; 21% of the moderate|moderate genotype frequency and 44% of the mild^CP^|severe genotype frequency.

#### Penetrance of c.5603A>T (p.(Asn1868Ile)): 5% to 65%

It is likely that incomplete penetrance occurs for several – if not all – known mild variants. However, in this study only the allele with the strongest evidence of incomplete penetrance was considered: the non-complex p.(Asn1868Ile). We implemented a penetrance of 5% when this allele is passed on from an unaffected carrier, as its penetrance is estimated at 5% in the general population.^9,20^ An alternative penetrance of 65% is implemented when a STGD1 patient passes p.(Asn1868Ile) to their child, which is the penetrance rate we observed among relatives in a study cohort of 27 families.^9,20^ For comparison, we included both a 65% and a 5% penetrance calculation when the p.(Asn1868Ile) variant was inherited from an unaffected *ABCA4* variant carrier. The higher penetrance rate should be considered when the carrier has first-degree relatives with STGD1, most often a sibling or a parent with STGD1.

## Results

### Composition of datasets

In total, data on *ABCA4* variants in 5,579 patients were collected (BAP dataset), consisting of 1,619 unique variants. Ethnicity data showed the following ancestries: non-Finnish European (67.8%), East Asian (9.7%), Latinx/Admixed American (7.0%), African (6.8%), Other (4.5%), South Asian (4.1%), Finnish (0.02%) and Ashkenazi Jewish (0.02%) (Supplemental Table 1). An EM-gnomAD dataset was constructed with corresponding ethnicity rates containing 132,890 alleles.

### Cumulative AF of severe, moderately severe and mild alleles

Of the 1,619 variants in the BAP dataset, 1,250 variants (77.2%) could be categorized into one or multiple severity groups (Table 1, Supplemental Table 2). 1,065 variants were categorized to a single severity group, the other 185 corresponded to more than one potential severity category. In total, 82 variants were categorized as benign (Supplemental Material and Supplemental Figure 1: categorization steps 3, 4 and 14). Furthermore, 66 null alleles from the EM-gnomAD dataset that were absent from the BAP dataset were added to the sum AF of severe variants. Finally, the sum AF of potential pathogenic alleles in the EM-gnomAD dataset was 7-8%.The total sum AFs per severity category per ethnic group can be found in Supplemental Table 3.

**Table 1.**
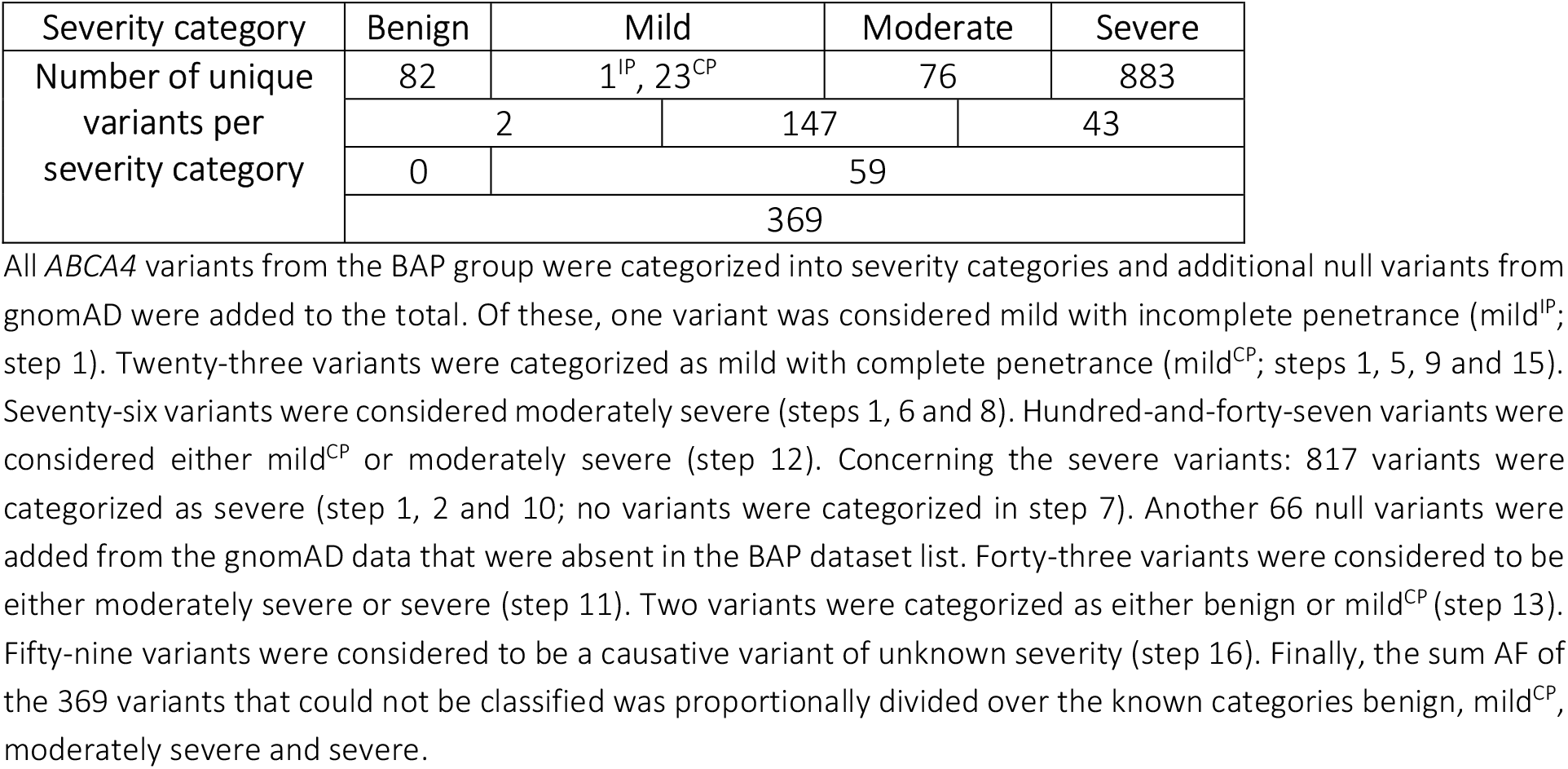
Variant categorization

### Genotype-based inheritance risk

The genotype frequencies per severity category in the general population based on the EM-gnomAD dataset are shown in Table 2. It shows that 5% of individuals carry at least one p.(Asn1868Ile) allele and 10% of individuals carry at least one (potentially) pathogenic *ABCA4* allele other than p.(Asn1868Ile). The inheritance risks for five genotype scenarios were calculated based on the EM-gnomAD dataset (Figure 1 and Supplemental Table 4). Results per gnomAD ethnic population can be found in Supplemental Figures 2-8.

The risk of STGD1 for the offspring of a STGD1 patient highly depends on the patient’s genotype (Figure 1A). As the combination of a severe *ABCA4* variant with any other pathogenic *ABCA4* variant will cause STGD1, the inheritance risk for offspring of a patient with two severe alleles and a non-tested unaffected individual is highest, estimated at 2.8%-3.1% (1 in 35-32) in our population. This is twice the risk of STGD1 for offspring of a patient who harbors a severe and a mild^CP^ allele: 1.6%-1.8% (1 in 62-57). Of important note: The risk that offspring will develop STGD1 already in childhood (0.68%-0.81%, 1 in 148-124, for a patient who harbors two severe variants) is estimated 2 to 4-fold lower than the risk that offspring will develop STGD1 at any moment in life (2.8%-3.1%, 1 in 35-32, for a patient who harbors two severe variants). This is because the genotype severe|mild – which typically does not lead to an onset in childhood – is most prominently present in any offspring genotype scenario.

**Table 2.** Estimates of *ABCA4* genotype frequencies in the general population

|  | Sev.<br>↓0.003944<br>↑0.004189 | Mod.<br>↓0.002920<br>↑0.003982 | Mild<sup>CP</sup><br>↓0.019127<br>↑0.020732 | N1868I<br>0.048694 | WT<br>↓0.925315<br>↑0.922404 |
| --- | --- | --- | --- | --- | --- |
| **Sev.**<br>↓0.003944<br>↑0.004189 | Sev.\|sev.<br>↓0.000016<br>↑0.000018 | Sev.\|mod.<br>↓2*0.000012<br>↑2*0.000017 | Sev.\|mild<sup>CP</sup><br>↓2*0.000075<br>↑2*0.000087 | Sev.\|N1868I<br>↓2*0.000192<br>↑2*0.000204 | Sev.\|WT<br>↓2*0.003650<br>↑2*0.003864 |
| **Mod.**<br>↓0.002920<br>↑0.003982 |  | Mod.\|mod.<br>↓0.000009<br>↑0.000016 | Mod.\|mild<sup>CP</sup><br>↓2*0.000056<br>↑2*0.000083 | Mod.\|N1868I<br>↓2*0.000142<br>↑2*0.000194 | Mod.\|WT<br>↓2*0.002702<br>↑2*0.003673 |
| **Mild<sup>CP</sup>**<br>↓0.019127<br>↑0.020732 |  |  | Mild<sup>CP</sup>\|mild<sup>CP</sup><br>↓0.000366<br>↑0.000430 | Mild<sup>CP</sup>\|N1868I<br>↓2*0.000931<br>↑2*0.001010 | Mild<sup>CP</sup>\|WT<br>↓2*0.017699<br>↑2*0.019123 |
| **N1868I**<br>0.048694 |  |  |  | N1868I\|N1868I<br>0.002371 | N1868I\|WT<br>↓2*0.045057<br>↑2*0.044915 |
| **WT**<br>↓0.925315<br>↑0.922404 |  |  |  |  | WT\|WT<br>↓0.856208<br>↑0.850829 |

Mild<sup>CP</sup>, mild *ABCA4* variant with complete penetrance; mod., moderately severe effect; N1868I, mild *ABCA4* variant with incomplete penetrance, c.5603A>T p.(Asn1868Ile); sev., severe effect; WT, wildtype.
Sum AFs are presented of different severity categories in the ethnicity matched-gnomAD dataset. Data include the sum AFs of causative variants of unknown severity divided over mild<sup>CP</sup>, moderately severe and severe, and variants with unknown pathogenicity divided over benign, mild<sup>CP</sup>, moderately severe and severe. ↓ represents the underestimate and ↑ the overestimate of the sum AFs and the corresponding genotype frequencies. Genotype frequencies are calculated using Hardy-Weinberg's principle, $p^2 + 2*p*q + q^2 = 1$ .
■ Genotype results in STGD1 phenotype; ■ Genotype may result in STGD1 phenotype; ■ Genotype is unlikely to result in a STGD1 phenotype but it might contribute to a pathogenic genotype in offspring; □ Genotype does not result in a STGD1 phenotype and does not contribute to a pathogenic genotype in offspring.

**Figure 1.**
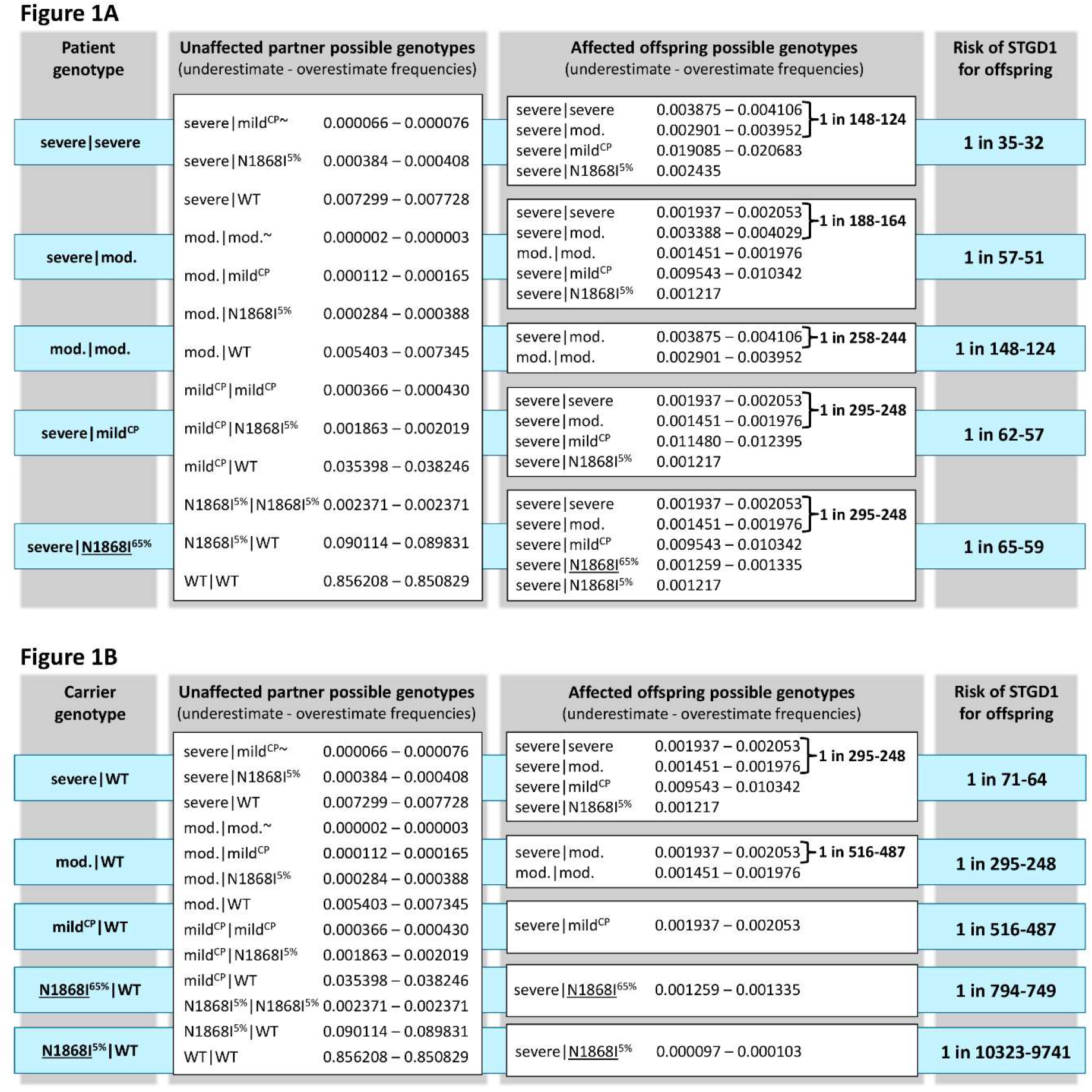
Genotype estimates for offspring of a STGD1 patient (A) or a carrier of a pathogenic *ABCA4* variant (B) and an unaffected partner with an unknown genotype. The blue boxes present the genotype scenarios for a STGD1 patient (A) and a known *ABCA4* variant carrier (B) who have a child with an unaffected individual harboring an unknown *ABCA4* genotype (white box on the left side). Ranges indicate the estimates based on the pathogenic sum allele frequency underestimate and overestimate in the ethnically matched-gnomAD. The risks are divided by genotype in the white boxes on the right side. The numbers in the blue boxes on the right represent the total risk of having affected offspring. Two different penetrance rates of p.(Asn1868Ile) (hereafter: N1868I) alleles were implemented: the N1868I allele in the general population has a penetrance of approximately 5%, whereas the N1868I allele within families with affected individuals shows higher penetrance, roughly estimated at 65%. The higher penetrance is therefore likely only applicable if the offspring inherited the N1868I allele from the affected parent (<u>N1868I</u> allele with underscore) or from the unaffected carrier of an *ABCA4* variant in whose (first degree) family that same allele had been penetrant. } specifies the risk of offspring with a severe|severe or severe|moderately severe genotype, who most likely manifest STGD1 in childhood. ∼ An individual with the severe|mild^CP^ or moderately severe|moderately severe genotype may be unaffected at the age they conceive a child and be diagnosed with STGD1 shortly or a long time thereafter. The presented risk estimates do not apply to situations where both (future) parents have STGD1, in which case it is generally feasibly to narrow down the possibly inheritance scenario’s based on known *ABCA4* genotypes of both (future) parents. Based on our Dutch patient data (E.H.R., personal communication), 44% of STGD1 patients carrying a severe|mild^CP^ genotype and 21% of patients with the moderate|moderate genotype received the diagnosis after the average age of European parents of children born in 2018 (32 years). The allele frequencies for these genotypes are corrected for those percentages. N1868I, mild p.(Asn1868Ile) *ABCA4* variant with incomplete penetrance. Mild^CP^, mild *ABCA4* variant with complete penetrance. Mod., *ABCA4* variant with moderately severe effect on ABCA4 function. WT, wildtype *ABCA4* allele.

The risks for offspring of unaffected *ABCA4* variant carriers varies tremendously, again the severity of the *ABCA4* allele being the most important determinant. When an *ABCA4* variant carrier from a STGD1 family (for instance, a child or genetically tested sibling of a patient) will have a child with an unaffected non-tested individual who does not have relatives with STGD1, the inheritance risk varies between 0.13% (1 in 794, for carriers of a mild^IP^ allel) and 1.6% (1 in 64, for carriers of a severe allele; Figure 1B). The risk that offspring develops STGD1 in childhood is estimated at practically zero for a known carrier of a mild allele to 0.34%-0.40% (1 in 295-248) for a known carrier of a severe allele.

## Discussion

Genetic counseling in STGD1 demands a personalized approach. Generalized counseling in autosomal recessive diseases based on the premise that a genotype consisting of two pathogenic alleles causes disease is inadequate in STGD1 because of the considerable genotypic and phenotypic variability. Two main aspects highlight the relevance of a personalized, genotype-directed counseling approach. First, the inheritance risk of STGD1 highly depends on the severity of the *ABCA4* alleles the parent carries. Accordingly, risks in our population were estimated to range 0.68%-3.1% (1 in 148-32), in case of one parent being a STGD1 patient, and 0.13-1.6% (1 in 794-64), in case of one parent being a known carrier of an *ABCA4* variant from a STGD1 family. As such, a sibling or child of a patient should be informed that, depending on the severity of the allele they carry, the inheritance risk of STGD1 at any moment in life for their children is either negligible or quite real. Second, people who seek counseling for family planning often involve a patient with an early onset of the disease. They might want to be counseled specifically for the risk that their offspring develops STGD1 at a young age. The inheritance risks for genotypes that generally lead to STGD1 in childhood are 2 to 4-fold lower than the total STGD1 inheritance risks.

These personalized risk estimates enable STGD1 patients and their family to make informed decisions regarding carrier testing of a healthy partner and/or reproductive decisions. Consequently, one may consider *ABCA4* testing of an unaffected partner. Today, whole-gene sequencing will reveal up to 95% of causal variants in clinically well-characterized STGD1 cases. If no putative causal *ABCA4* variants are identified in the partner, the risk of STGD1 for the offspring accordingly will decrease up to 20-fold. When a (likely) causal variant is found, prenatal or pre-implantation genetic diagnostics or prenatal testing could be discussed with the prospective parents, taking into account knowledge of the pathogenicity as well as the severity of the variant.

Our study provides data on variant severity for many variants in the *ABCA4* gene. This data can help clinicians interpret the consequences of a specific *ABCA4* variant for their patient only if the robustness of the data underlying the variant categorization is carefully considered. It is likely that a proportion of variants, allocated on the basis of lesser robust data, have been assigned to the wrong severity category. However, the methods we used did not create a bias of variant allocation into one category over the other, and thus provide an adequate estimate of the overall sum AF per severity category. Due to insufficient data, 428/1,619 unique *ABCA4* variants could not be assigned to one specific or either of two severity categories. Their total AF in the EM-gnomAD dataset was only 0.29%. Excluding these variants from the analysis completely would only be justified if each of these was benign, which is unlikely. Finally, stop-gain variants may not always lead to a null allele, but of all the stop-gain variants that had a low severity odds ratio, none was found homozygously less often than expected in the BAP dataset, indicating that they are moderately severe or severe. Genetic counseling for individuals carrying a variant that was assigned a severity category on the basis of the lesser robust data, should especially consider that the inheritance risk lies on a range of estimates provided by several possibly relevant genotypic scenarios.

STGD1 inheritance risks are greatly influenced by differences in penetrance, due to the high frequency of the incompletely penetrant p.(Asn1868Ile) allele. The estimated penetrance rates of 5% for the general population to 65% for familial cases are based on the only data available to date: population data and data of a small number of STGD1 families, respectively.^9,20^ When p.(Asn1868Ile) is inherited from an unaffected 1^st^ degree relative of a STGD1 case, the penetrance might be lower than 65% as also the total number of genetic modifiers outside the *ABCA4* gene may be lower, as compared to the situation where the p.(Asn1868Ile) allele has been penetrant in a first degree relative. We recommend counselors to carefully consider these potential differences in penetrance on the basis of the family history of both (future) parents.

When estimating the risk of STGD1 for the offspring of STGD1 cases or unaffected carriers of *ABCA4* alleles, several other factors need to be considered. First, given the hypothesized polygenic or multifactorial nature of STGD1 for a significant fraction of cases (∼25%), the risk estimates may vary depending on the culture, population and country. Second, *ABCA4* carriership reported in literature varies tremendously (6-20%),^17,31,32^ and accordingly the chances of meeting an unaffected partner who carries a (potentially) pathogenic *ABCA4* allele may vary. This variability is highly influenced by genetic testing method, variant interpretation, new insights over time, and possibly by population. In our study, the AF of (potentially) pathogenic alleles totals 7-8%, meaning that 14-15% of individuals in the general population carries at least one potentially pathogenic *ABCA4* allele.

We calculated risks on the basis of the AFs in different ethnic subpopulations, which in theory would result in a more precise, personalized estimate. Risks in subpopulations differed up to 230% from the general population that was ethnically matched to the patient group. However, risk estimates for separate ethnicities should be considered with special caution. First and most importantly, genetic variation within populations is likely much larger than between populations.^33^ Moreover, one’s self-reported ethnicity might not correspond with ethnicity grouping on the basis of genetic similarities. Local AFs can highly impact the risk assessment, e.g. c.5882G>A is extremely frequent in the Somali population, potentially increasing the risk of STGD1 in this population.^34^ Local AFs may have to be incorporated into the risk assessment when they differ highly from the published data. Another source of uncertainty concerns insufficient knowledge on complex alleles, especially in understudied populations. Inheritance risk is overestimated when unrecognized complex alleles are included in sum AFs in duplicate. Furthermore, the causal variants as well as the severity of variants are less studied especially in populations outside Europe and North America. For some populations both the STGD1 patient group as well as the control group are relatively small or non-existing. Consequently, causal variants in these populations may be underreported and unidentified, leading to an underestimation of the total risk in these populations. Finally, prevalence studies in understudied populations might help indicate if the risk of STGD1 and of passing on STGD1 to offspring indeed varies across populations or whether there still remain a lot of disease variants to be identified.

The tremendous advances in genetic testing and knowledge of the *ABCA4* gene made over the past decade should now be followed by the improvement of genetic counseling to STGD1 patients and their families. In the future, inheritance risk estimations need to be optimized on the basis of new studies on the functional effect of *ABCA4* variants and increased knowledge on genotype frequencies worldwide as well as differences between populations and within populations, haplotypes, allele severity, and penetrance. For patients and their families today, the presented genotype-directed approach can help inform them about their personalized inheritance risks more reliably than generalized autosomal recessive risk counseling.

## Data Availability

Patient data have been uploaded to the LOVD ABCA4 database.

http://www.lovd.nl/abca4

## Acknowledgements

We would like to acknowledge Rona Jualla van Oudenhoven for her advice on using inclusive language.

## Ethics approval

Not applicable.

## Contributorship Statement

SSC performed variant analyses. EHR performed the inheritance risk calculations. MB and ZC provided pre-existing experimental variant information. EdB, SR, LHW, CMD, ATVvS and FPMC provided expert input. FPMC supervised the project. SSC, EHR and FPMC designed the study. SSC, EHR, ATVvS and FPMC wrote the manuscript. All authors reviewed and approved the manuscript.

## Supplementary Material

Supplemental Figure 1. Flowchart variant severity allocation. Fully and shaded grey squared show the number corresponding to the classification step it represents. Fully grey squares indicate the more robust classifications while shaded squares indicate lesser robust classifications. Brackets indicate the number of variants that have been categorized in that step.

Supplemental Figure 2A and B. Genotype estimates for offspring of a STGD1 patient (A) or a carrier of a pathogenic *ABCA4* variant (B) and an unaffected partner with an unknown genotype in the African population.

Supplemental Figure 3A and B. Genotype estimates for offspring of a STGD1 patient (A) or a carrier of a pathogenic *ABCA4* variant (B) and an unaffected partner with an unknown genotype in the Latino/Admixed American population.

Supplemental Figure 4A and B. Genotype estimates for offspring of a STGD1 patient (A) or a carrier of a pathogenic *ABCA4* variant (B) and an unaffected partner with an unknown genotype in the Ashkenazi Jewish population.

Supplemental Figure 5A and B. Genotype estimates for offspring of a STGD1 patient (A) or a carrier of a pathogenic *ABCA4* variant (B) and an unaffected partner with an unknown genotype in the East Asian population.

Supplemental Figure 6A and B. Genotype estimates for offspring of a STGD1 patient (A) or a carrier of a pathogenic *ABCA4* variant (B) and an unaffected partner with an unknown genotype in the Finnish European population.

Supplemental Figure 7A and B. Genotype estimates for offspring of a STGD1 patient (A) or a carrier of a pathogenic *ABCA4* variant (B) and an unaffected partner with an unknown genotype in the non-Finnish European population.

Supplemental Figure 8A and B. Genotype estimates for offspring of a STGD1 patient (A) or a carrier of a pathogenic *ABCA4* variant (B) and an unaffected partner with an unknown genotype in the South Asian population.

Supplemental Table 1. Ethnic makeup of the BAP based control group

Supplemental Table 2. Data analysis for pathogenicity and severity allocation (i.e. allele frequency test, homozygosity test and occurrence with mild/severe ratio test).

Supplemental Table 3. The allele frequencies of severe, moderately severe, and mild *ABCA4* variants per gnomAD population.

Supplemental Table 4. Calculation sheet for the risk of STGD1 for offspring of STGD1 patients or known *ABCA4* variant carriers

