## Supplemental Figures for "Genetic risk estimates for offspring of patients with Stargardt disease"

### Supplemental Figure 1

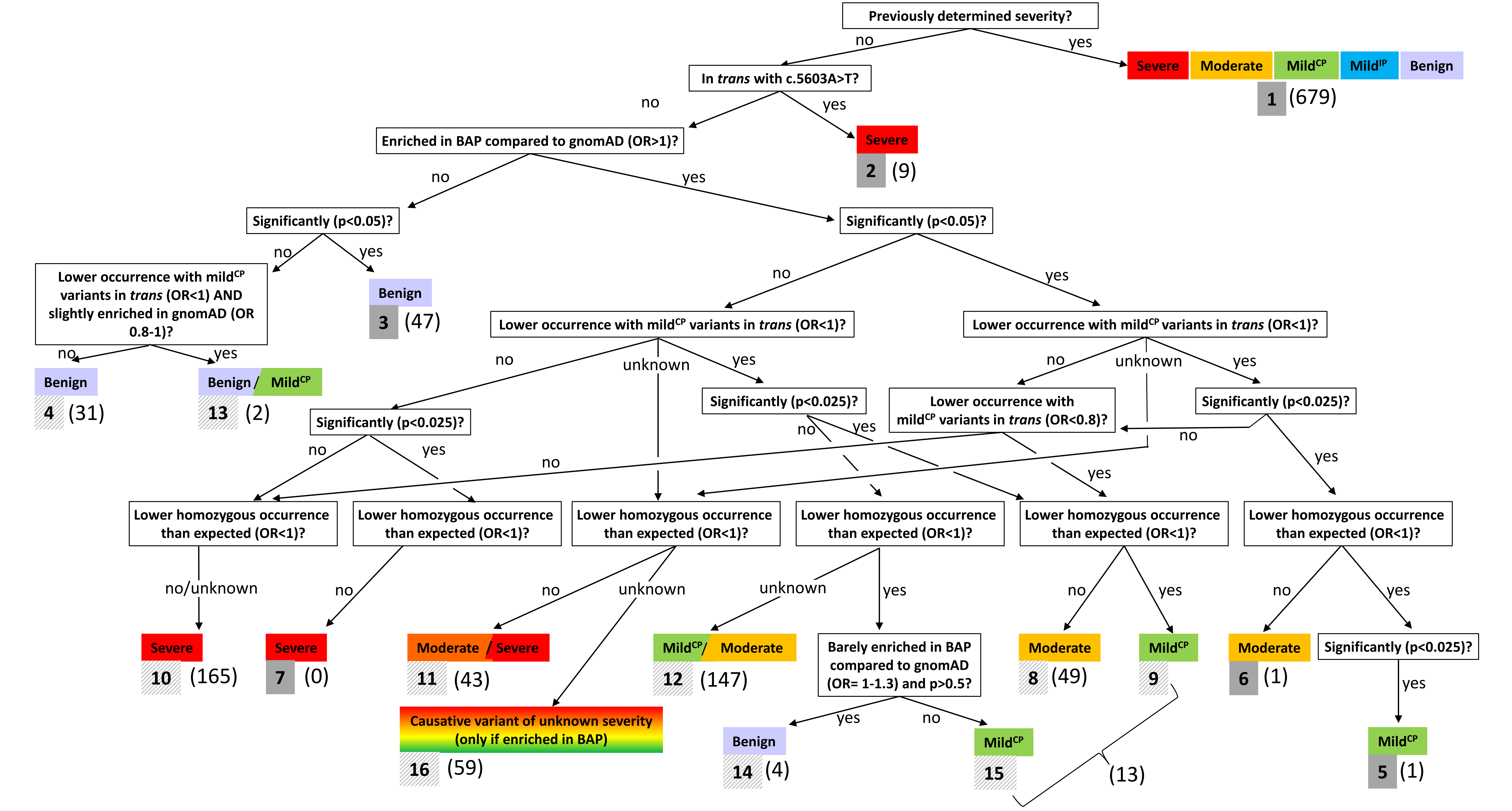

**Supplemental Figure 2A.** Genotype estimates for offspring of a STGD1 patient and an unaffected partner with an unknown genotype in the **African population**

**Unaffected partner possible genotypes** (underestimate - overestimate frequencies)

| Genotype | Frequency |
|---|---|
| severe\|mild$^{CP}$~ | 0.000245 – 0.000316 |
| severe\|N1868I$^{5\%}$ | 0.000096 – 0.000128 |
| severe\|WT | 0.009244 – 0.012310 |
| mod.\|mod.~ | 0.000004 – 0.000004 |
| mod.\|mild$^{CP}$ | 0.000458 – 0.000491 |
| mod.\|N1868I$^{5\%}$ | 0.000079 – 0.000087 |
| mod.\|WT | 0.007601 – 0.008423 |
| mild$^{CP}$\|mild$^{CP}$ | 0.003110 – 0.002908 |
| mild$^{CP}$\|N1868I$^{5\%}$ | 0.001072 – 0.001036 |
| mild$^{CP}$\|WT | 0.103230 – 0.099785 |
| N1868I$^{5\%}$\|N1868I$^{5\%}$ | 0.000092 – 0.000092 |
| N1868I$^{5\%}$\|WT | 0.017789 – 0.017884 |
| WT\|WT | 0.856590 – 0.856113 |

| Patient genotype | Affected offspring possible genotypes (underestimate - overestimate frequencies) | } | Risk of STGD1 for offspring |
|---|---|---|---|
| **severe\|severe** | severe\|severe 0.004792 – 0.006377<br>severe\|mod. 0.004072 – 0.004505<br>severe\|mild$^{CP}$ 0.055613 – 0.053722<br>severe\|N1868I$^{5\%}$ 0.000481 | **1 in 113-92** | **1 in 15** |
| **severe\|mod.** | severe\|severe 0.002396 – 0.003188<br>severe\|mod. 0.004432 – 0.005441<br>mod.\|mod. 0.002036 – 0.002253<br>severe\|mild$^{CP}$ 0.027806 – 0.026861<br>severe\|N1868I$^{5\%}$ 0.000240 | **1 in 146-116** | **1 in 27-26** |
| **mod.\|mod.** | severe\|mod. 0.004792 – 0.006377<br>mod.\|mod. 0.004072 – 0.004505 | **1 in 209-157** | **1 in 113-92** |
| **severe\|mild$^{CP}$** | severe\|severe 0.002396 – 0.003188<br>severe\|mod. 0.002036 – 0.002253<br>severe\|mild$^{CP}$ 0.030202 – 0.030049<br>severe\|N1868I$^{5\%}$ 0.000240 | **1 in 226-184** | **1 in 29-28** |
| **severe\|<u>N1868I</u>$^{65\%}$** | severe\|severe 0.002396 – 0.003188<br>severe\|mod. 0.002036 – 0.002253<br>severe\|mild$^{CP}$ 0.027806 – 0.026861<br>severe\|<u>N1868I</u>$^{65\%}$ 0.001558 – 0.002072<br>severe\|N1868I$^{5\%}$ 0.000240 | **1 in 226-184** | **1 in 29** |

**Supplemental Figure 2A: Genotype estimates for offspring of a STGD1 patient and an unaffected partner with an unknown genotype in the African population.**

**Supplemental Figure 2B.** Genotype estimates for offspring of a carrier of a pathogenic *ABCA4* variant and an unaffected partner with an unknown genotype in the **African population**

**Unaffected partner possible genotypes** (underestimate - overestimate frequencies)

| | |
|---|---|
| severe\|mild$^{CP}$~ | 0.000245 – 0.000316 |
| severe\|N1868I$^{5\%}$ | 0.000096 – 0.000128 |
| severe\|WT | 0.009244 – 0.012310 |
| mod.\|mod.~ | 0.000004 – 0.000004 |
| mod.\|mild$^{CP}$ | 0.000458 – 0.000491 |
| mod.\|N1868I$^{5\%}$ | 0.000079 – 0.000087 |
| mod.\|WT | 0.007601 – 0.008423 |
| mild$^{CP}$\|mild$^{CP}$ | 0.003110 – 0.002908 |
| mild$^{CP}$\|N1868I$^{5\%}$ | 0.001072 – 0.001036 |
| mild$^{CP}$\|WT | 0.103230 – 0.099785 |
| N1868I$^{5\%}$\|N1868I$^{5\%}$ | 0.000092 – 0.000092 |
| N1868I$^{5\%}$\|WT | 0.017789 – 0.017784 |
| WT\|WT | 0.856590 – 0.856113 |

| Carrier genotype | Affected offspring possible genotypes (underestimate - overestimate frequencies) | | | Risk of STGD1 for offspring |
|---|---|---|---|---|
| **severe\|WT** | severe\|severe | 0.002396 – 0.003188 | } **1 in 226-184** | **1 in 31** |
| | severe\|mod. | 0.002036 – 0.002253 | } | |
| | severe\|mild$^{CP}$ | 0.027806 – 0.026861 | | |
| | severe\|N1868I$^{5\%}$ | 0.000240 | | |
| **mod.\|WT** | severe\|mod. | 0.002396 – 0.000316 | } **1 in 417-314** | **1 in 226-184** |
| | mod.\|mod. | 0.002036 – 0.002253 | | |
| **mild$^{CP}$\|WT** | severe\|mild$^{CP}$ | 0.002396 – 0.003188 | | **1 in 417-314** |
| **N1868I$^{65\%}$\|WT** | severe\|N1868I$^{65\%}$ | 0.001558 – 0.002072 | | **1 in 642-483** |
| **N1868I$^{5\%}$\|WT** | severe\|N1868I$^{5\%}$ | 0.000120 – 0.000159 | | **1 in 8347-6273** |

**Supplemental Figure 2B: Genotype estimates for offspring of a carrier of a pathogenic *ABCA4* variant and an unaffected partner with an unknown genotype in the African population.**

**Supplemental Figure 3A.** Genotype estimates for offspring of a STGD1 patient and an unaffected partner with an unknown genotype in the **Latino/Admixed American population**

**Unaffected partner possible genotypes** (underestimate - overestimate frequencies)

| Genotype | Frequency |
|---|---|
| severe\|mild$^{CP}$~ | 0.000032 – 0.000035 |
| severe\|N1868I$^{5\%}$ | 0.000100 – 0.000127 |
| severe\|WT | 0.004325 – 0.005464 |
| mod.\|mod.~ | 0.000008 – 0.000013 |
| mod.\|mild$^{CP}$ | 0.000191 – 0.000218 |
| mod.\|N1868I$^{5\%}$ | 0.000265 – 0.000348 |
| mod.\|WT | 0.011426 – 0.014988 |
| mild$^{CP}$\|mild$^{CP}$ | 0.000253 – 0.000192 |
| mild$^{CP}$\|N1868I$^{5\%}$ | 0.000703 – 0.000612 |
| mild$^{CP}$\|WT | 0.030343 – 0.026394 |
| N1868I$^{5\%}$\|N1868I$^{5\%}$ | 0.000489 – 0.000498 |
| N1868I$^{5\%}$\|WT | 0.042172 – 0.042154 |
| WT\|WT | 0.909593 – 0.908821 |

| Patient genotype | Affected offspring possible genotypes (underestimate - overestimate frequencies) | | } | Risk of STGD1 for offspring |
|---|---|---|---|---|
| **severe\|severe** | severe\|severe | 0.002229 – 0.002813 | **1 in 122-94** | **1 in 40-39** |
| | severe\|mod. | 0.005984 – 0.007790 | | |
| | severe\|mild$^{CP}$ | 0.015887 – 0.013821 | | |
| | severe\|N1868I$^{5\%}$ | 0.001105 | | |
| **severe\|mod.** | severe\|severe | 0.001114 – 0.001406 | **1 in 192-149** | **1 in 60-55** |
| | severe\|mod. | 0.004088 – 0.005301 | | |
| | mod.\|mod. | 0.002974 – 0.003895 | | |
| | severe\|mild$^{CP}$ | 0.007944 – 0.006911 | | |
| | severe\|N1868I$^{5\%}$ | 0.000553 | | |
| **mod.\|mod.** | severe\|mod. | 0.002229 – 0.002813 | **1 in 449-356** | **1 in 122-94** |
| | mod.\|mod. | 0.005948 – 0.007790 | | |
| **severe\|mild$^{CP}$** | severe\|severe | 0.001114 – 0.001406 | **1 in 245-189** | **1 in 73-71** |
| | severe\|mod. | 0.002974 – 0.003895 | | |
| | severe\|mild$^{CP}$ | 0.009058 – 0.008317 | | |
| | severe\|N1868I$^{5\%}$ | 0.000553 | | |
| **severe\|<u>N1868I</u>$^{65\%}$** | severe\|severe | 0.001114 – 0.001406 | **1 in 245-189** | **1 in 75-73** |
| | severe\|mod. | 0.002974 – 0.003895 | | |
| | severe\|mild$^{CP}$ | 0.007944 – 0.006911 | | |
| | severe\|<u>N1868I</u>$^{65\%}$ | 0.000724 – 0.000914 | | |
| | severe\|N1868I$^{5\%}$ | 0.000553 | | |

**Supplemental Figure 3A. Genotype estimates for offspring of a STGD1 patient and an unaffected partner with an unknown genotype in the Latino/Admixed American population**

**Supplemental Figure 3B.** Genotype estimates for offspring of a carrier of a pathogenic *ABCA4* variant and an unaffected partner with an unknown genotype in the **Latino/Admixed American population**

| Unaffected partner possible genotypes (underestimate - overestimate frequencies) | |
|---|---|
| severe\|mild$^{CP}$~ | 0.000032 – 0.000035 |
| severe\|N1868I$^{5\%}$ | 0.000100 – 0.000127 |
| severe\|WT | 0.004325 – 0.005464 |
| mod.\|mod.~ | 0.000008 – 0.000013 |
| mod.\|mild$^{CP}$ | 0.000191 – 0.000218 |
| mod.\|N1868I$^{5\%}$ | 0.000265 – 0.000348 |
| mod.\|WT | 0.011426 – 0.014988 |
| mild$^{CP}$\|mild$^{CP}$ | 0.000253 – 0.000192 |
| mild$^{CP}$\|N1868I$^{5\%}$ | 0.000703 – 0.000612 |
| mild$^{CP}$\|WT | 0.030343 – 0.026394 |
| N1868I$^{5\%}$\|N1868I$^{5\%}$ | 0.000489 – 0.000498 |
| N1868I$^{5\%}$\|WT | 0.042172 – 0.042154 |
| WT\|WT | 0.909593 – 0.908821 |

| Carrier genotype | Affected offspring possible genotypes (underestimate - overestimate frequencies) | | | Risk of STGD1 for offspring |
|---|---|---|---|---|
| **severe\|WT** | severe\|severe | 0.001114 – 0.001406 | } **1 in 245-189** | **1 in 79-78** |
| | severe\|mod. | 0.002974 – 0.003895 | } | |
| | severe\|mild$^{CP}$ | 0.007794 – 0.006911 | | |
| | severe\|N1868I$^{5\%}$ | 0.000553 | | |
| **mod.\|WT** | severe\|mod. | 0.001114 – 0.001406 | } **1 in 897-711** | **1 in 245-189** |
| | mod.\|mod. | 0.002974 – 0.003895 | | |
| **mild$^{CP}$\|WT** | severe\|mild$^{CP}$ | 0.001114 – 0.001406 | | **1 in 897-711** |
| **N1868I$^{65\%}$\|WT** | severe\|N1868I$^{65\%}$ | 0.000724 – 0.000914 | | **1 in 1381-1094** |
| **N1868I$^{5\%}$\|WT** | severe\|N1868I$^{5\%}$ | 0.000056 – 0.000070 | | **1 in 17498-14221** |

**Supplemental Figure 3B: Genotype estimates for offspring of a carrier of a pathogenic *ABCA4* variant and an unaffected partner with an unknown genotype in the Latino/Admixed American population.**

**Supplemental Figure 4A.** Genotype estimates for offspring of a STGD1 patient and an unaffected partner with an unknown genotype in the **Askenazi Jewish population**

**Unaffected partner possible genotypes** (underestimate - overestimate frequencies)

| | |
|---|---|
| severe\|mild$^{CP}$~ | 0.000040 – 0.000041 |
| severe\|N1868I$^{5\%}$ | 0.000061 – 0.000061 |
| severe\|WT | 0.001610 – 0.001604 |
| mod.\|mod.~ | 0.000007 – 0.000007 |
| mod.\|mild$^{CP}$ | 0.000573 – 0.000601 |
| mod.\|N1868I$^{5\%}$ | 0.000384 – 0.000391 |
| mod.\|WT | 0.010173 – 0.010334 |
| mild$^{CP}$\|mild$^{CP}$ | 0.002615 – 0.002781 |
| mild$^{CP}$\|N1868I$^{5\%}$ | 0.003507 – 0.003617 |
| mild$^{CP}$\|WT | 0.092892 – 0.095594 |
| N1868I$^{5\%}$\|N1868I$^{5\%}$ | 0.001176 – 0.001176 |
| N1868I$^{5\%}$\|WT | 0.062278 – 0.062162 |
| WT\|WT | 0.824618 – 0.821544 |

| Patient genotype | Affected offspring possible genotypes (underestimate - overestimate frequencies) | | } | Risk of STGD1 for offspring |
|---|---|---|---|---|
| **severe\|severe** | severe\|severe | 0.000855 – 0.000853 | **1 in 156-153** | **1 in 17-16** |
| | severe\|mod. | 0.005572 – 0.005670 | | |
| | severe\|mild$^{CP}$ | 0.051111 – 0.052707 | | |
| | severe\|N1868I$^{5\%}$ | 0.001715 | | |
| **severe\|mod.** | severe\|severe | 0.000428 – 0.000426 | **1 in 275-271** | **1 in 30** |
| | severe\|mod. | 0.003214 – 0.003261 | | |
| | mod.\|mod. | 0.002786 – 0.002835 | | |
| | severe\|mild$^{CP}$ | 0.002555 – 0.026354 | | |
| | severe\|N1868I$^{5\%}$ | 0.000857 | | |
| **mod.\|mod.** | severe\|mod. | 0.000855 – 0.000853 | **1 in 1173-1169** | **1 in 156-153** |
| | mod.\|mod. | 0.005572 – 0.005670 | | |
| **severe\|mild$^{CP}$** | severe\|severe | 0.000428 – 0.000426 | **1 in 311-307** | **1 in 33-32** |
| | severe\|mod. | 0.002768 – 0.002835 | | |
| | severe\|mild$^{CP}$ | 0.025983 – 0.026780 | | |
| | severe\|N1868I$^{5\%}$ | 0.000857 | | |
| **severe\|<u>N1868I</u>$^{65\%}$** | severe\|severe | 0.000428 – 0.000426 | **1 in 311-307** | **1 in 33** |
| | severe\|mod. | 0.002786 – 0.002825 | | |
| | severe\|mild$^{CP}$ | 0.025555 – 0.026354 | | |
| | severe\|<u>N1868I</u>$^{65\%}$ | 0.000278 – 0.000277 | | |
| | severe\|N1868I$^{5\%}$ | 0.000857 | | |

**Supplemental Figure 4A: Genotype estimates for offspring of a STGD1 patient and an unaffected partner with an unknown genotype in the Askenazi Jewish population.**
The blue boxes present the genotype scenarios for a STGD1 patient who haves a child with an unaffected individual harboring an unknown *ABCA4* genotype (white box on the left side). Ranges indicate the estimates based on the pathogenic sum allele frequency underestimate and overestimate in the Askenazi Jewish population. The risks are divided by genotype in the white boxes on the right side. The numbers in the blue boxes on the right represent the total risk of having affected offspring. Two different penetrance rates of p.(Asn1868Ile) (hereafter: N1868I) alleles were implemented: the N1868I allele in the general population has a penetrance of approximately 5%, whereas the N1868I allele within families with affected individuals shows higher penetrance, roughly estimated at 65%. The higher penetrance is therefore likely only applicable if the offspring inherited the N1868I allele from the affected parent (<u>N1868I</u> allele with underscore) or from the unaffected carrier of an *ABCA4* variant in whose (first degree) family that same allele had been penetrant.
} specifies the risk of offspring with a severe|severe or severe|moderately severe genotype, who most likely manifest STGD1 in childhood.
~ An individual with the severe|mild$^{CP}$ or moderately severe|moderately severe genotype may be unaffected at the age they conceive a child and be diagnosed with STGD1 shortly or a long time thereafter. The presented risk estimates do not apply to situations where both (future) parents have STGD1, in which case it is generally feasibly to narrow down the possibly inheritance scenarios based on known *ABCA4* genotypes of both (future) parents. Based on our Dutch patient data (E.H.R., personal communication), 44% of STGD1 patients carrying a severe|mild$^{CP}$ genotype and 21% of patients with the moderately severe|moderately severe genotype received the diagnosis after the average age of European parents of children born in 2018 (32 years). The allele frequencies for these genotypes are corrected for those percentages.
N1868I, mild p.(Asn1868Ile) *ABCA4* variant with incomplete penetrance. Mild$^{CP}$, mild *ABCA4* variant with complete penetrance. Mod., *ABCA4* variant with moderately severe effect on ABCA4 function. WT, wildtype *ABCA4* allele.

**Supplemental Figure 4B.** Genotype estimates for offspring of a carrier of a pathogenic *ABCA4* variant and an unaffected partner with an unknown genotype in the **Askenazi Jewish population**

**Unaffected partner possible genotypes** (underestimate - overestimate frequencies)

| Genotype | Frequency |
|---|---|
| severe\|mild$^{CP}$~ | 0.000040 – 0.000041 |
| severe\|N1868I$^{5\%}$ | 0.000061 – 0.000061 |
| severe\|WT | 0.001610 – 0.001604 |
| mod.\|mod.~ | 0.000007 – 0.000007 |
| mod.\|mild$^{CP}$ | 0.000573 – 0.000601 |
| mod.\|N1868I$^{5\%}$ | 0.000384 – 0.000391 |
| mod.\|WT | 0.010173 – 0.010334 |
| mild$^{CP}$\|mild$^{CP}$ | 0.002615 – 0.002781 |
| mild$^{CP}$\|N1868I$^{5\%}$ | 0.003507 – 0.003617 |
| mild$^{CP}$\|WT | 0.092892 – 0.095594 |
| N1868I$^{5\%}$\|N1868I$^{5\%}$ | 0.001176 – 0.001176 |
| N1868I$^{5\%}$\|WT | 0.062278 – 0.062162 |
| WT\|WT | 0.824618 – 0.821544 |

| Carrier genotype | Affected offspring possible genotypes (underestimate - overestimate frequencies) | | | Risk of STGD1 for offspring |
|---|---|---|---|---|
| **severe\|WT** | severe\|severe | 0.000428 – 0.000426 | } **1 in 311-307** | **1 in 34-33** |
| | severe\|mod. | 0.002786 – 0.002835 | } | |
| | severe\|mild$^{CP}$ | 0.025555 – 0.026354 | | |
| | severe\|N1868I$^{5\%}$ | 0.000857 | | |
| **mod.\|WT** | severe\|mod. | 0.000428 – 0.000426 | } **1 in 2345-2338** | **1 in 311-307** |
| | mod.\|mod. | 0.002786 – 0.002835 | | |
| **mild$^{CP}$\|WT** | severe\|mild$^{CP}$ | 0.000428 – 0.000426 | | **1 in 2345-2338** |
| **<u>N1868I</u>$^{65\%}$\|WT** | severe\|<u>N1868I</u>$^{65\%}$ | 0.000278 – 0.000277 | | **1 in 3608-3597** |
| **<u>N1868I</u>$^{5\%}$\|WT** | severe\|<u>N1868I</u>$^{5\%}$ | 0.000021 – 0.000021 | | **1 in 46901-46759** |

**Supplemental Figure 4B: Genotype estimates for offspring of a carrier of a pathogenic *ABCA4* variant and an unaffected partner with an unknown genotype in the Askenazi Jewish population.**

The blue boxes present the genotype scenarios for a known carrier of an *ABCA4* variant who haves a child with an unaffected individual harboring an unknown *ABCA4* genotype (white box on the left side). Ranges indicate the estimates based on the pathogenic sum allele frequency underestimate and overestimate in the Askenazi Jewish population. The risks are divided by genotype in the white boxes on the right side. The numbers in the blue boxes on the right represent the total risk of having affected offspring. Two different penetrance rates of p.(Asn1868Ile) (hereafter: N1868I) alleles were implemented: the N1868I allele in the general population has a penetrance of approximately 5%, whereas the N1868I allele within families with affected individuals shows higher penetrance, roughly estimated at 65%. The higher penetrance is therefore likely only applicable if the offspring inherited the N1868I allele from the affected parent (<u>N1868I</u> allele with underscore) or from the unaffected carrier of an *ABCA4* variant in whose (first degree) family that same allele had been penetrant.

} specifies the risk of offspring with a severe|severe or severe|moderately severe genotype, who most likely manifest STGD1 in childhood.

~ An individual with the severe|mild$^{CP}$ or moderately severe|moderately severe genotype may be unaffected at the age they conceive a child and be diagnosed with STGD1 shortly or a long time thereafter. The presented risk estimates do not apply to situations where both (future) parents have STGD1, in which case it is generally feasibly to narrow down the possibly inheritance scenarios based on known *ABCA4* genotypes of both (future) parents. Based on our Dutch patient data (E.H.R., personal communication), 44% of STGD1 patients carrying a severe|mild$^{CP}$ genotype and 21% of patients with the moderately severe|moderately severe genotype received the diagnosis after the average age of European parents of children born in 2018 (32 years). The allele frequencies for these genotypes are corrected for those percentages.

N1868I, mild p.(Asn1868Ile) *ABCA4* variant with incomplete penetrance. Mild$^{CP}$, mild *ABCA4* variant with complete penetrance. Mod., *ABCA4* variant with moderately severe effect on ABCA4 function. WT, wildtype *ABCA4* allele.

**Supplemental Figure 5A.** Genotype estimates for offspring of a STGD1 patient and an unaffected partner with an unknown genotype in the **East Asian population**

**Unaffected partner possible genotypes** (underestimate - overestimate frequencies)

| Genotype | Frequency |
|---|---|
| severe\|mild$^{CP}$~ | 0.000049 – 0.000039 |
| severe\|N1868I$^{5\%}$ | 0.000000 – 0.000000 |
| severe\|WT | 0.006891 – 0.007036 |
| mod.\|mod.~ | 0.000001 – 0.000005 |
| mod.\|mild$^{CP}$ | 0.000057 – 0.000125 |
| mod.\|N1868I$^{5\%}$ | 0.000000 – 0.000000 |
| mod.\|WT | 0.003569 – 0.009905 |
| mild$^{CP}$\|mild$^{CP}$ | 0.000247 – 0.000154 |
| mild$^{CP}$\|N1868I$^{5\%}$ | 0.000000 – 0.000000 |
| mild$^{CP}$\|WT | 0.030761 – 0.024279 |
| N1868I$^{5\%}$\|N1868I$^{5\%}$ | 0.000000 – 0.000000 |
| N1868I$^{5\%}$\|WT | 0.000000 – 0.000000 |
| WT\|WT | 0.958336 – 0.958336 |

| Patient genotype | Affected offspring possible genotypes (underestimate - overestimate frequencies) | } | Risk of STGD1 for offspring |
|---|---|---|---|
| **severe\|severe** | severe\|severe 0.003470 – 0.003538<br>severe\|mod. 0.001814 – 0.005021<br>severe\|mild$^{CP}$ 0.015680 – 0.012376<br>severe\|N1868I$^{5\%}$ 0.000000 | **1 in 189-117** | **1 in 48** |
| **severe\|mod.** | severe\|severe 0.001735 – 0.001769<br>severe\|mod. 0.002642 – 0.004279<br>mod.\|mod. 0.000907 – 0.002510<br>severe\|mild$^{CP}$ 0.007840 – 0.006188<br>severe\|N1868I$^{5\%}$ 0.000000 | **1 in 228-165** | **1 in 76-68** |
| **mod.\|mod.** | severe\|mod. 0.003470 – 0.003538<br>mod.\|mod. 0.001814 – 0.005021 | **1 in 288-283** | **1 in 189-117** |
| **severe\|mild$^{CP}$** | severe\|severe 0.001735 – 0.001769<br>severe\|mod. 0.000907 – 0.002510<br>severe\|mild$^{CP}$ 0.009575 – 0.007957<br>severe\|N1868I$^{5\%}$ 0.000000 | **1 in 379-234** | **1 in 82** |
| **severe\|<u>N1868I</u>$^{65\%}$** | severe\|severe 0.001735 – 0.001769<br>severe\|mod. 0.000907 – 0.002510<br>severe\|mild$^{CP}$ 0.007840 – 0.006188<br>severe\|<u>N1868I</u>$^{65\%}$ 0.001128 – 0.001150<br>severe\|N1868I$^{5\%}$ 0.000000 | **1 in 379-234** | **1 in 86** |

**Supplemental Figure 5A: Genotype estimates for offspring of a STGD1 patient and an unaffected partner with an unknown genotype in the East Asian population.**

**Supplemental Figure 5B.** Genotype estimates for offspring of a carrier of a pathogenic *ABCA4* variant and an unaffected partner with an unknown genotype in the **East Asian population**

**Unaffected partner possible genotypes** (underestimate - overestimate frequencies)

| Genotype | Frequency |
|---|---|
| severe\|mild$^{CP}$~ | 0.000049 – 0.000039 |
| severe\|N1868I$^{5\%}$ | 0.000000 – 0.000000 |
| severe\|WT | 0.006891 – 0.007036 |
| mod.\|mod.~ | 0.000001 – 0.000005 |
| mod.\|mild$^{CP}$ | 0.000057 – 0.000125 |
| mod.\|N1868I$^{5\%}$ | 0.000000 – 0.000000 |
| mod.\|WT | 0.003569 – 0.009905 |
| mild$^{CP}$\|mild$^{CP}$ | 0.000247 – 0.000154 |
| mild$^{CP}$\|N1868I$^{5\%}$ | 0.000000 – 0.000000 |
| mild$^{CP}$\|WT | 0.030761 – 0.024279 |
| N1868I$^{5\%}$\|N1868I$^{5\%}$ | 0.000000 – 0.000000 |
| N1868I$^{5\%}$\|WT | 0.000000 – 0.000000 |
| WT\|WT | 0.958336 – 0.958336 |

| Carrier genotype | Affected offspring possible genotypes (underestimate - overestimate frequencies) | | | Risk of STGD1 for offspring |
|---|---|---|---|---|
| **severe\|WT** | severe\|severe | 0.001735 – 0.001769 | } **1 in 379-234** | **1 in 95-96** |
| | severe\|mod. | 0.000907 – 0.002510 | } | |
| | severe\|mild$^{CP}$ | 0.007840 – 0.006188 | | |
| | severe\|N1868I$^{5\%}$ | 0.000000 | | |
| **mod.\|WT** | severe\|mod. | 0.001735 – 0.001769 | } **1 in 576-565** | **1 in 379-234** |
| | mod.\|mod. | 0.000907 – 0.002510 | | |
| **mild$^{CP}$\|WT** | severe\|mild$^{CP}$ | 0.001735 – 0.001769 | | **1 in 576-565** |
| **<u>N1868I</u>$^{65\%}$\|WT** | severe\|<u>N1868I</u>$^{65\%}$ | 0.001128 – 0.001150 | | **1 in 887-870** |
| **<u>N1868I</u>$^{5\%}$\|WT** | severe\|<u>N1868I</u>$^{5\%}$ | 0.000087 – 0.000088 | | **1 in 11528-11307** |

**Supplemental Figure 5B: Genotype estimates for offspring a carrier of a pathogenic *ABCA4* variant and an unaffected partner with an unknown genotype in the East Asian population.**

**Supplemental Figure 6A.** Genotype estimates for offspring of a STGD1 patient and an unaffected partner with an unknown genotype in the **Finnish European population**

**Unaffected partner possible genotypes** (underestimate - overestimate frequencies)

| Genotype | Frequency |
|---|---|
| severe\|mild$^{CP}$~ | 0.000028 – 0.000047 |
| severe\|N1868I$^{5\%}$ | 0.000123 – 0.000122 |
| severe\|WT | 0.002926 – 0.002862 |
| mod.\|mod.~ | 0.000000 – 0.000000 |
| mod.\|mild$^{CP}$ | 0.000017 – 0.000035 |
| mod.\|N1868I$^{5\%}$ | 0.000033 – 0.000040 |
| mod.\|WT | 0.000793 – 0.000949 |
| mild$^{CP}$\|mild$^{CP}$ | 0.000403 – 0.001195 |
| mild$^{CP}$\|N1868I$^{5\%}$ | 0.001579 – 0.002719 |
| mild$^{CP}$\|WT | 0.037685 – 0.063881 |
| N1868I$^{5\%}$\|N1868I$^{5\%}$ | 0.001547 – 0.001547 |
| N1868I$^{5\%}$\|WT | 0.073825 – 0.072679 |
| WT\|WT | 0.881003 – 0.853860 |

| Patient genotype | Affected offspring possible genotypes (underestimate - overestimate frequencies) | | } | Risk of STGD1 for offspring |
|---|---|---|---|---|
| **severe\|severe** | severe\|severe | 0.001538 – 0.001516 | **1 in 510-493** | **1 in 42-26** |
| | severe\|mod. | 0.000422 – 0.000512 | | |
| | severe\|mild$^{CP}$ | 0.020057 – 0.034536 | | |
| | severe\|N1868I$^{5\%}$ | 0.001966 | | |
| **severe\|mod.** | severe\|severe | 0.000769 – 0.000758 | **1 in 572-564** | **1 in 77-49** |
| | severe\|mod. | 0.000980 – 0.001014 | | |
| | mod.\|mod. | 0.000211 – 0.000256 | | |
| | severe\|mild$^{CP}$ | 0.010029 – 0.017268 | | |
| | severe\|N1868I$^{5\%}$ | 0.000983 | | |
| **mod.\|mod.** | severe\|mod. | 0.001538 – 0.001516 | **1 in 660-650** | **1 in 510-493** |
| | mod.\|mod. | 0.000422 – 0.000512 | | |
| **severe\|mild$^{CP}$** | severe\|severe | 0.000769 – 0.000758 | **1 in 1021-986** | **1 in 78-50** |
| | severe\|mod. | 0.000211 – 0.000256 | | |
| | severe\|mild$^{CP}$ | 0.010798 – 0.018026 | | |
| | severe\|N1868I$^{5\%}$ | 0.000983 | | |
| **severe\|<u>N1868I</u>$^{65\%}$** | severe\|severe | 0.000769 – 0.000758 | **1 in 1021-986** | **1 in 80-51** |
| | severe\|mod. | 0.000211 – 0.000256 | | |
| | severe\|mild$^{CP}$ | 0.010029 – 0.017268 | | |
| | severe\|<u>N1868I</u>$^{65\%}$ | 0.000500 – 0.000493 | | |
| | severe\|N1868I$^{5\%}$ | 0.000983 | | |

**Supplemental Figure 6A: Genotype estimates for offspring of a STGD1 patient and an unaffected partner with an unknown genotype in the Finnish European population.**
The blue boxes present the genotype scenarios for a STGD1 patient who haves a child with an unaffected individual harboring an unknown *ABCA4* genotype (white box on the left side). Ranges indicate the estimates based on the pathogenic sum allele frequency underestimate and overestimate in the Finnish European population. The risks are divided by genotype in the white boxes on the right side. The numbers in the blue boxes on the right represent the total risk of having affected offspring. Two different penetrance rates of p.(Asn1868Ile) (hereafter: N1868I) alleles were implemented: the N1868I allele in the general population has a penetrance of approximately 5%, whereas the N1868I allele within families with affected individuals shows higher penetrance, roughly estimated at 65%. The higher penetrance is therefore likely only applicable if the offspring inherited the N1868I allele from the affected parent (<u>N1868I</u> allele with underscore) or from the unaffected carrier of an *ABCA4* variant in whose (first degree) family that same allele had been penetrant.
} specifies the risk of offspring with a severe|severe or severe|moderately severe genotype, who most likely manifest STGD1 in childhood.
~ An individual with the severe|mild$^{CP}$ or moderately severe|moderately severe genotype may be unaffected at the age they conceive a child and be diagnosed with STGD1 shortly or a long time thereafter. The presented risk estimates do not apply to situations where both (future) parents have STGD1, in which case it is generally feasibly to narrow down the possibly inheritance scenarios based on known *ABCA4* genotypes of both (future) parents. Based on our Dutch patient data (E.H.R., personal communication), 44% of STGD1 patients carrying a severe|mild$^{CP}$ genotype and 21% of patients with the moderately severe|moderately severe genotype received the diagnosis after the average age of European parents of children born in 2018 (32 years). The allele frequencies for these genotypes are corrected for those percentages.
N1868I, mild p.(Asn1868Ile) *ABCA4* variant with incomplete penetrance. Mild$^{CP}$, mild *ABCA4* variant with complete penetrance. Mod., *ABCA4* variant with moderately severe effect on ABCA4 function. WT, wildtype *ABCA4* allele.

**Supplemental Figure 6B.** Genotype estimates for offspring of a carrier of a pathogenic *ABCA4* variant and an unaffected partner with an unknown genotype in the **Finnish European population**

<table>
<tr><th colspan="2">Unaffected partner possible genotypes (underestimate - overestimate frequencies)</th></tr>
<tr><td>severe|mild$^{CP}$~</td><td>0.000028 – 0.000047</td></tr>
<tr><td>severe|N1868I$^{5\%}$</td><td>0.000123 – 0.000122</td></tr>
<tr><td>severe|WT</td><td>0.002926 – 0.002862</td></tr>
<tr><td>mod.|mod.~</td><td>0.000000 – 0.000000</td></tr>
<tr><td>mod.|mild$^{CP}$</td><td>0.000017 – 0.000035</td></tr>
<tr><td>mod.|N1868I$^{5\%}$</td><td>0.000033 – 0.000040</td></tr>
<tr><td>mod.|WT</td><td>0.000793 – 0.000949</td></tr>
<tr><td>mild$^{CP}$|mild$^{CP}$</td><td>0.000403 – 0.001195</td></tr>
<tr><td>mild$^{CP}$|N1868I$^{5\%}$</td><td>0.001579 – 0.002719</td></tr>
<tr><td>mild$^{CP}$|WT</td><td>0.037685 – 0.063881</td></tr>
<tr><td>N1868I$^{5\%}$|N1868I$^{5\%}$</td><td>0.001547 – 0.001547</td></tr>
<tr><td>N1868I$^{5\%}$|WT</td><td>0.073825 – 0.072679</td></tr>
<tr><td>WT|WT</td><td>0.881003 – 0.853860</td></tr>
</table>

<table>
<tr><th>Carrier genotype</th><th colspan="3">Affected offspring possible genotypes (underestimate - overestimate frequencies)</th><th>Risk of STGD1 for offspring</th></tr>
<tr><td rowspan="4">**severe|WT**</td><td>severe|severe</td><td>0.000769 – 0.000758</td><td rowspan="2">} **1 in 1021-986**</td><td rowspan="4">**1 in 83-52**</td></tr>
<tr><td>severe|mod.</td><td>0.000211 – 0.000256</td></tr>
<tr><td>severe|mild$^{CP}$</td><td>0.010029 – 0.017268</td><td></td></tr>
<tr><td>severe|N1868I$^{5\%}$</td><td>0.000983</td><td></td></tr>
<tr><td rowspan="2">**mod.|WT**</td><td>severe|mod.</td><td>0.000769 – 0.000758</td><td>} **1 in 1320-1300**</td><td rowspan="2">**1 in 1021-986**</td></tr>
<tr><td>mod.|mod.</td><td>0.000211 – 0.000256</td><td></td></tr>
<tr><td>**mild$^{CP}$|WT**</td><td>severe|mild$^{CP}$</td><td>0.000769 – 0.000759</td><td></td><td>**1 in 1320-1300**</td></tr>
<tr><td>**N1868I$^{65\%}$|WT**</td><td>severe|N1868I$^{65\%}$</td><td>0.000500 – 0.002072</td><td></td><td>**1 in 2030-2000**</td></tr>
<tr><td>**N1868I$^{5\%}$|WT**</td><td>severe|N1868I$^{5\%}$</td><td>0.000038 – 0.000038</td><td></td><td>**1 in 26390-26005**</td></tr>
</table>

**Supplemental Figure 7A.** Genotype estimates for offspring of a STGD1 patient and an unaffected individual with an unknown genotype in the **non-Finnish European population**

**Unaffected partner possible genotypes** (underestimate - overestimate frequencies)

| Genotype | Frequency |
|---|---|
| severe\|mild$^{CP}$~ | 0.000065 – 0.000079 |
| severe\|N1868I$^{5\%}$ | 0.000561 – 0.000579 |
| severe\|WT | 0.007973 – 0.008179 |
| mod.\|mod.~ | 0.000002 – 0.000003 |
| mod.\|mild$^{CP}$ | 0.000097 – 0.000148 |
| mod.\|N1868I$^{5\%}$ | 0.000368 – 0.000479 |
| mod.\|WT | 0.005219 – 0.006772 |
| mild$^{CP}$\|mild$^{CP}$ | 0.000286 – 0.000395 |
| mild$^{CP}$\|N1868I$^{5\%}$ | 0.002170 – 0.002551 |
| mild$^{CP}$\|WT | 0.030809 – 0.036064 |
| N1868I$^{5\%}$\|N1868I$^{5\%}$ | 0.004122 – 0.004122 |
| N1868I$^{5\%}$\|WT | 0.117061 – 0.116552 |
| WT\|WT | 0.831135 – 0.823914 |

| Patient genotype | Affected offspring possible genotypes (underestimate - overestimate frequencies) | | Risk of STGD1 for offspring |
|---|---|---|---|
| **severe\|severe** | severe\|severe | 0.004300 – 0.004418 } **1 in 140-123** | **1 in 37-32** |
| | severe\|mod. | 0.002843 – 0.003703 } | |
| | severe\|mild$^{CP}$ | 0.016856 – 0.019815 | |
| | severe\|N1868I$^{5\%}$ | 0.003210 | |
| **severe\|mod.** | severe\|severe | 0.002150 – 0.002209 } **1 in 175-160** | **1 in 58-51** |
| | severe\|mod. | 0.003572 – 0.004060 } | |
| | mod.\|mod. | 0.001422 – 0.001851 | |
| | severe\|mild$^{CP}$ | 0.008428 – 0.009908 | |
| | severe\|N1868I$^{5\%}$ | 0.001605 | |
| **mod.\|mod.** | severe\|mod. | 0.004300 – 0.004418 } **1 in 233-226** | **1 in 140-123** |
| | mod.\|mod. | 0.002843 – 0.003703 | |
| **severe\|mild$^{CP}$** | severe\|severe | 0.002150 – 0.002209 } **1 in 280-246** | **1 in 63-56** |
| | severe\|mod. | 0.001422 – 0.001851 } | |
| | severe\|mild$^{CP}$ | 0.010578 – 0.012117 | |
| | severe\|N1868I$^{5\%}$ | 0.001605 | |
| **severe\|N1868I$^{65\%}$** | severe\|severe | 0.002150 – 0.002209 } **1 in 280-246** | **1 in 67-59** |
| | severe\|mod. | 0.001422 – 0.001851 } | |
| | severe\|mild$^{CP}$ | 0.008428 – 0.009908 | |
| | severe\|N1868I$^{65\%}$ | 0.001397 – 0.001436 | |
| | severe\|N1868I$^{5\%}$ | 0.001605 | |

**Supplemental Figure 7A: Genotype estimates for offspring of a STGD1 patient and an unaffected partner with an unknown genotype in the non-Finnish European population.**

**Supplemental Figure 7B.** Genotype estimates for offspring of a carrier of a pathogenic *ABCA4* variant and an unaffected partner with an unknown genotype in the **non-Finnish European population**

**Unaffected partner possible genotypes** (underestimate - overestimate frequencies)

| Genotype | Frequency |
|---|---|
| severe\|mild$^{CP}$~ | 0.000065 – 0.000079 |
| severe\|N1868I$^{5\%}$ | 0.000561 – 0.000579 |
| severe\|WT | 0.007973 – 0.008179 |
| mod.\|mod.~ | 0.000002 – 0.000003 |
| mod.\|mild$^{CP}$ | 0.000097 – 0.000148 |
| mod.\|N1868I$^{5\%}$ | 0.000368 – 0.000479 |
| mod.\|WT | 0.005219 – 0.006772 |
| mild$^{CP}$\|mild$^{CP}$ | 0.000286 – 0.000395 |
| mild$^{CP}$\|N1868I$^{5\%}$ | 0.002170 – 0.002551 |
| mild$^{CP}$\|WT | 0.030809 – 0.036064 |
| N1868I$^{5\%}$\|N1868I$^{5\%}$ | 0.004122 – 0.004122 |
| N1868I$^{5\%}$\|WT | 0.117061 – 0.116552 |
| WT\|WT | 0.831135 – 0.823914 |

| Carrier genotype | Affected offspring possible genotypes (underestimate - overestimate frequencies) | Risk of STGD1 for offspring |
|---|---|---|
| **severe\|WT** | severe\|severe 0.002150 – 0.002209 }<br>severe\|mod. 0.001422 – 0.001851 } **1 in 280-246**<br>severe\|mild$^{CP}$ 0.008428 – 0.009908<br>severe\|N1868I$^{5\%}$ 0.001605 | **1 in 74-64** |
| **mod.\|WT** | severe\|mod. 0.002150 – 0.002209 } **1 in 465-453**<br>mod.\|mod. 0.001422 – 0.001851 | **1 in 280-246** |
| **mild$^{CP}$\|WT** | severe\|mild$^{CP}$ 0.002150 – 0.002209 | **1 in 465-453** |
| **N1868I$^{65\%}$\|WT** | severe\|N1868I$^{65\%}$ 0.001397 – 0.001436 | **1 in 716-696** |
| **N1868I$^{5\%}$\|WT** | severe\|N1868I$^{5\%}$ 0.000107 – 0.000110 | **1 in 9303-9054** |

**Supplemental Figure 7B: Genotype estimates for offspring of a carrier of a pathogenic *ABCA4* variant and an unaffected partner with an unknown genotype in the non-Finnish European population.**

**Supplemental Figure 8A.** Genotype estimates for offspring of a STGD1 patient and an unaffected individual with an unknown genotype in the **South Asian population**

**Unaffected partner possible genotypes** (underestimate - overestimate frequencies)

| Genotype | Frequency |
|---|---|
| severe\|mild$^{CP}$~ | 0.000062 – 0.000063 |
| severe\|N1868I$^{5\%}$ | 0.000118 – 0.000124 |
| severe\|WT | 0.006232 – 0.006562 |
| mod.\|mod.~ | 0.000000 – 0.000001 |
| mod.\|mild$^{CP}$ | 0.000040 – 0.000081 |
| mod.\|N1868I$^{5\%}$ | 0.000033 – 0.000071 |
| mod.\|WT | 0.001739 – 0.003744 |
| mild$^{CP}$\|mild$^{CP}$ | 0.000473 – 0.000431 |
| mild$^{CP}$\|N1868I$^{5\%}$ | 0.000784 – 0.000748 |
| mild$^{CP}$\|WT | 0.041577 – 0.039675 |
| N1868I$^{5\%}$\|N1868I$^{5\%}$ | 0.000325 – 0.000325 |
| N1868I$^{5\%}$\|WT | 0.034468 – 0.034460 |
| WT\|WT | 0.914052 – 0.913607 |

| Patient genotype | Affected offspring possible genotypes (underestimate - overestimate frequencies) | | Risk of STGD1 for offspring |
|---|---|---|---|
| **severe\|severe** | severe\|severe | 0.003206 – 0.003374 } **1 in 243-188** | **1 in 37** |
| | severe\|mod. | 0.000906 – 0.001949 } | |
| | severe\|mild$^{CP}$ | 0.021704 – 0.020714 | |
| | severe\|N1868I$^{5\%}$ | 0.000901 | |
| **severe\|mod.** | severe\|severe | 0.001603 – 0.001687 } **1 in 273-230** | **1 in 65-62** |
| | severe\|mod. | 0.002056 – 0.002662 } | |
| | mod.\|mod. | 0.000453 – 0.000974 | |
| | severe\|mild$^{CP}$ | 0.010852 – 0.010357 | |
| | severe\|N1868I$^{5\%}$ | 0.000451 | |
| **mod.\|mod.** | severe\|mod. | 0.003206 – 0.003374 } **1 in 312-296** | **1 in 243-188** |
| | mod.\|mod. | 0.000906 – 0.001949 | |
| **severe\|mild$^{CP}$** | severe\|severe | 0.001603 – 0.001687 } **1 in 486-376** | **1 in 67-66** |
| | severe\|mod. | 0.000453 – 0.000974 } | |
| | severe\|mild$^{CP}$ | 0.012455 – 0.012044 | |
| | severe\|N1868I$^{5\%}$ | 0.000451 | |
| **severe\|<u>N1868I</u>$^{65\%}$** | severe\|severe | 0.001603 – 0.001687 } **1 in 486-376** | **1 in 69** |
| | severe\|mod. | 0.000453 – 0.000974 } | |
| | severe\|mild$^{CP}$ | 0.010852 – 0.010357 | |
| | severe\|<u>N1868I</u>$^{65\%}$ | 0.001042 – 0.001097 | |
| | severe\|N1868I$^{5\%}$ | 0.000451 | |

**Supplemental Figure 8A: Genotype estimates for offspring of a STGD1 patient and an unaffected partner with an unknown genotype in the South Asian population.**

**Supplemental Figure 8B.** Genotype estimates for offspring of a carrier of a pathogenic *ABCA4* variant and an unaffected partner with an unknown genotype in the **South Asian population**

**Unaffected partner possible genotypes** (underestimate - overestimate frequencies)

| Genotype | Frequency |
|---|---|
| severe\|mild$^{CP}$~ | 0.000062 – 0.000063 |
| severe\|N1868I$^{5\%}$ | 0.000118 – 0.000124 |
| severe\|WT | 0.006232 – 0.006562 |
| mod.\|mod.~ | 0.000000 – 0.000001 |
| mod.\|mild$^{CP}$ | 0.000040 – 0.000081 |
| mod.\|N1868I$^{5\%}$ | 0.000033 – 0.000071 |
| mod.\|WT | 0.001739 – 0.003744 |
| mild$^{CP}$\|mild$^{CP}$ | 0.000473 – 0.000431 |
| mild$^{CP}$\|N1868I$^{5\%}$ | 0.000784 – 0.000748 |
| mild$^{CP}$\|WT | 0.041577 – 0.039675 |
| N1868I$^{5\%}$\|N1868I$^{5\%}$ | 0.000325 – 0.000325 |
| N1868I$^{5\%}$\|WT | 0.034468 – 0.034460 |
| WT\|WT | 0.914052 – 0.913607 |

| Carrier genotype | Affected offspring possible genotypes (underestimate - overestimate frequencies) | | | Risk of STGD1 for offspring |
|---|---|---|---|---|
| **severe\|WT** | severe\|severe | 0.001603 – 0.001687 | } **1 in 486-376** | **1 in 75-74** |
| | severe\|mod. | 0.000453 – 0.000974 | } | |
| | severe\|mild$^{CP}$ | 0.027806 – 0.026861 | | |
| | severe\|N1868I$^{5\%}$ | 0.000451 | | |
| **mod.\|WT** | severe\|mod. | 0.001603 – 0.001687 | } **1 in 624-593** | **1 in 486-376** |
| | mod.\|mod. | 0.000453 – 0.000974 | | |
| **mild$^{CP}$\|WT** | severe\|mild$^{CP}$ | 0.001603 – 0.001687 | | **1 in 624-593** |
| **N1868I$^{65\%}$\|WT** | severe\|N1868I$^{65\%}$ | 0.001042 – 0.001097 | | **1 in 960-912** |
| **N1868I$^{5\%}$\|WT** | severe\|N1868I$^{5\%}$ | 0.000080 – 0.000084 | | **1 in 12476-11854** |

**Supplemental Figure 8B: Genotype estimates for offspring of a carrier of a pathogenic *ABCA4* variant and an unaffected partner with an unknown genotype in the South Asian population.**
The blue boxes present the genotype scenarios for a known carrier of an *ABCA4* variant who haves a child with an unaffected individual harboring an unknown *ABCA4* genotype (white box on the left side). Ranges indicate the estimates based on the pathogenic sum allele frequency underestimate and overestimate in the South Asian population. The risks are divided by genotype in the white boxes on the right side. The numbers in the blue boxes on the right represent the total risk of having affected offspring. Two different penetrance rates of p.(Asn1868Ile) (hereafter: N1868I) alleles were implemented: the N1868I allele in the general population has a penetrance of approximately 5%, whereas the N1868I allele within families with affected individuals shows higher penetrance, roughly estimated at 65%. The higher penetrance is therefore likely only applicable if the offspring inherited the N1868I allele from the affected parent (N1868I allele with underscore) or from the unaffected carrier of an *ABCA4* variant in whose (first degree) family that same allele had been penetrant.
} specifies the risk of offspring with a severe|severe or severe|moderately severe genotype, who most likely manifest STGD1 in childhood.
~ An individual with the severe|mild$^{CP}$ or moderately severe|moderately severe genotype may be unaffected at the age they conceive a child and be diagnosed with STGD1 shortly or a long time thereafter. The presented risk estimates do not apply to situations where both (future) parents have STGD1, in which case it is generally feasibly to narrow down the possibly inheritance scenarios based on known *ABCA4* genotypes of both (future) parents. Based on our Dutch patient data (E.H.R., personal communication), 44% of STGD1 patients carrying a severe|mild$^{CP}$ genotype and 21% of patients with the moderately severe|moderately severe genotype received the diagnosis after the average age of European parents of children born in 2018 (32 years). The allele frequencies for these genotypes are corrected for those percentages.
N1868I, mild p.(Asn1868Ile) *ABCA4* variant with incomplete penetrance. Mild$^{CP}$, mild *ABCA4* variant with complete penetrance. Mod., *ABCA4* variant with moderately severe effect on ABCA4 function. WT, wildtype *ABCA4* allele.
