## Supplemental Materials and Methods for "Genetic risk estimates for offspring of patients with Stargardt disease"

**Supplemental Material and Methods**

Variant data collection

We collected all the papers that were published until 2020 that contain *ABCA4* variants in patients with an autosomal recessive retinal dystrophy by searching the following terms in PubMed:

(ABCA4[All Fields] OR (("Stargardt disease"[All Fields] OR "Macula Lutea"[All Fields]) AND ("Genetics"[All Fields] OR "mutation"[All Fields] OR "Sequence Analysis"[All Fields] OR "gene panel"[TiAb]))) OR ("Retinal Dystrophies"[All Fields] AND ("mutation"[All Fields] OR "Sequence Analysis"[All Fields] AND "gene panel"[TiAb])) (Cornelis et al, in preparation).^27^

Creating an ethnically matched control group

*ABCA4* allele frequency data from the general population were taken from the Genome Aggregation Database (gnomAD), downloaded on 13 April 2021. We aimed to take the largest gnomAD sample that ethnically matches our bi-allelic patient (BAP) dataset as best as possible. Of note: In gnomAD, ethnicity categories have been created by principal component analyses, grouping together those genetic data most alike. This approach likely does not match the patient-reported ethnicity, which is the most common means of ethnicity collected in medicine. We therefore used one combined “general population” allele frequency that was matched to the ethnicities reported in published *ABCA4* data, which likely mainly represents the populations of Europe and North-America.

We ethnically matched the gnomAD allele data to the BAP data, taking the following steps:

First, we adjusted the published ethnicities from the BAP dataset data to the gnomAD populations: Black and people of African ancestry 🡪 African; Latinx, Hispanic, people of ancestry of Latin American countries 🡪 Latino/Admixed American; Ashkenazi Jewish 🡪 Ashkenazi Jewish; Chinese, Japanese, Mongolian, North Korean, South Korean, Taiwanese and people of East Asian ancestry 🡪 East Asian; Finnish 🡪 Finnish European; White and people of European ancestry 🡪 non-Finnish European; Afghan, people from Pakistan ancestry, Indian, Nepalese, people from Bhutan ancestry, Bangladeshi, Sri Lankan and people of South Asian ancestry 🡪 South Asian; People with mixed or other reported ethnicities 🡪 Other.

Second, approximately half of the studies that did not report ethnicities were published in the USA. We therefore assigned ethnicity proportions in the USA population, as reported by https://statisticalatlas.com/United-States/Race-and-Ethnicity, to 50% of the data in which no ethnicity was reported. The other 50% of data with unknown ethnicities originated from European papers and were assigned according to English and Welsh ethnicity data (https://www.ethnicity-facts-figures.service.gov.uk/uk-population-by-ethnicity/national-and-regional-populations/population-of-england-and-wales/latest) by lack of more detailed data, due to many European laws on recording ethnic data (https://www.researchgate.net/publication/233314223_Collecting_ethnic_statistics_in_Europe_A_review).

Third, per gnomAD population we calculated the average allele number from exonic and splice site variants – as annotated in gnomAD - to create the largest possible ethnically matched gnomAD (EM-gnomAD) dataset (Suppl. Table 1): To find the gnomAD ethnic population size that limits and thereby determines the total size of the EM-dataset, we calculated the ratio between BAP group % / gnomAD group % per ethnic population. The ratio of the ‘Other’ ethnic group was the largest and therefore determined the size of the EM-dataset: had the ‘Other’ group been larger in gnomAD, the EM-gnomAD could have been larger as well. We therefore took the size (average exonic allele number (AN)) of the ‘Other’ group in gnomAD as a reference size for the EM-gnomAD dataset:

∑ BAP(gnomAD Other AN)* (BAP dataset population group %) / (BAP dataset Other %)

We multiplied the original gnomAD AN of the ‘Other’ category with each of the ethnicity groups percentages in the BAP dataset and divided these by the Other group percentage in the BAP dataset. The resulting numbers of each ethnic group were then summed up to get to the total EM-gnomAD control AN. In this way the total adjusted gnomAD allele number became smaller than the actual gnomAD dataset but we maintained the trustworthiness of the smaller ‘Other’ group size from gnomAD, as this will affect the Fisher Exact tests that were used.

Severe variants test: previously established severity of variants

The following variants were defined as ‘previously established mild’, based on their reduced occurrence in homozygous configuration among patients, i.e. c.2588G>C, c.3113C>T, c.5882G>A, and c.6089G>A,^27^ or based on their behavior in genotype-phenotype correlations: c.769-784C>T and c.4253+43G>A,^5,18^ c.5603A>T,^9,17^ and/or based on expression levels c.6320G>A.^30^ To increase the robustness of this test, we defined variants ‘previously established severe’ only if they most likely lead to no effective ABCA4 activity. Therefore, severe variants in this analysis only included frameshift, stop-gain and canonical splice site variants as well as noncanonical splice site variants that were shown to lead to ≤5% of the WT product *in vitro*, the 5% being a conservative cut-off (Z. Corradi and F.P.M. Cremers, personal communication).^18,21-25,31-34^

Severity category assignment

We assigned variants to the categories ‘benign’, ‘mild^IP^’, ‘mild^CP^’, ‘moderately severe’ and ‘severe’ using the steps described below, each represented as well in a flow chart in Supplemental Figure 1. Of note, this categorization is meant to create a sum AF per severity category and not to categorize individual variants robustly. For severity allocation of individual variants, steps 1-3 and 5-7 make use of Fisher-Exact p-values <0.05 or <0.025 and are therefore more robust than steps 4 and 8-15. However, no correction for multiple testing has been performed and therefore the results should be appreciated with caution.

1. Variants with a previously determined severity. Established likely severe variants have been listed in previous studies: stop-gain, frameshift, canonical splice site variants, in addition to non-canonical splice site (NCSS) and deep-intronic (DI) variants that result in ≤25% normal RNA.^17,19-24,8,183,28-31^ Variants that were previously functionally or clinically determined to be mild, were assigned ‘Mild-complete penetrance’ (‘Mild^CP^’).^17,19-23,25,28-31^ NCSS and DI variants that are known to lead to >25-<70% of wild type RNA levels were assigned ‘Moderately severe’ and NCSS and DI variants known to lead to >70% and <85% of wild type RNA product were assigned ‘Mild^CP^’. Recently, the effect of a small selection of missense variants was assessed using expression levels, ATPase activity and genotype-phenotype correlations.^27^ Severity of variants was assigned accordingly.
2. The frequent c.5603A>T (p.(Asn1868Ile)) variant appears to have such a mild effect that it only causes STGD1 when in *trans* with a severe allele. As also suggested by others,^16^ this p.(Asn1868Ile) allele can serve as a litmus test to assign variants in *trans* (on the other allele) the status ‘severe’.^4,32^ Variants that were found in *trans* with the non-complex p.(Asn1868Ile) variant (i.e. p.(Asn1868Ile) without additionally potentially pathogenic variants on the same allele) in patients (confirmed by segregation analysis or by homozygous configuration) were assigned ‘**severe**’.^8,16,23^
3. AF test OR<1, p<0.05: **benign**
4. AF test OR<1: **benign**
5. AF test OR>1, p<0.05 AND homozygosity test OR<1, p<0.025 AND severity OR<1, p<0.025: **mild^CP^**
6. Homozygosity test OR≥1 AND severity OR<1, p<0.025: **moderately severe**
7. Homozygosity test OR≥1 AND severity OR>1, p<0.025: **severe** (no variants were categorized this way)
8. Homozygosity test OR≥1 AND severity OR<0.8: **moderately severe**
9. AF test OR>1, p<0.05 AND homozygosity test OR<1 AND severity OR<0.8: **mild^CP^**
10. Homozygosity test OR≥1 (also if the expected occurrence was 0) AND severity OR>1: **severe**
11. (Homozygosity test OR≥1 AND no data available for the severity OR,) OR (Homozygosity test OR≥1 AND severity OR>0.8): **moderately severe/severe**
12. AF test OR>1 AND variants do not occur homozygously and aren’t expected to occur homozygously AND severity OR <0.8: **mild^CP^/moderately severe**
13. AF test OR: >0.75, <1 AND severity OR<1: **benign/mild^CP^**
14. AF test OR >1, <1.3 that have a p-value of >0.5: **benign**
15. AF test OR>1.3 AND homozygosity test OR<1: **mild^CP^**
16. AF test OR>1, p<0.05 AND variants do not occur homozygously and are not expected to occur homozygously AND variants do not occur with known mild or known severe variants *in trans* (i.e. the severity OR test could not be performed): **VUS but likely causative**
17. The sum allele frequency of all the variants that could not be categorized as either of the aforementioned categories, was divided into those categories according to the ratio of sum allele frequencies of the categorized variants.
